# The potential crosstalk genes andmolecular mechanismsbetweenintracranial aneurysm and periodontitis

**DOI:** 10.1101/2023.05.18.23290206

**Authors:** Jian-huang Huang, Yao Chen, Yuan-bao Kang, Zheng-jian Yao, Jian-hua Song

**Affiliations:** Department of Neurosurgery, Affiliated Hospital of Putian University, Putian, Fujian Province, China

**Keywords:** Intracranial aneurysms, Periodontitis, crosstalk genes, immune infiltration, bioinformatics analysis

## Abstract

**Background:** The risk of intracranial aneurysm (IAs) development and rupture is significantly higher in patients with periodontitis (PD), suggesting an association between the two. However, the specific mechanisms of association between these two diseases have not been fully investigated. The aim of this study was to investigate potential crosstalk genes, pathways between IAs and PD.

**Methods:** We downloaded IAs and PD data from the Gene Expression Omnibus (GEO). Differentially expressed genes (DEGs) were identified, and functional enrichment analysis was performed on these overlapping DEGs. The protein-protein interaction (PPI) network was constructed and the cytoHubba plugin was used to identify PPI key genes. Next, weighted gene co-expression network analysis (WGCNA) was performed. Subsequently, we validated the key crosstalk genes in 2 independent external datasets. In addition, the immune cell landscape was assessed and the correlation of key crosstalk genes with each immune cell was calculated. Finally, transcription factors (TFs) regulating key crosstalk genes were explored and their expression levels were further validated in the full dataset.

**Results:** 127 overlapping DEGs were identified and functional enrichment analysis highlighted the important role of immune reflection in the pathogenesis. We identified ITGAX and COL4A2 as key crosstalk genes and the expression levels of them were significantly elevated in the test dataset and external validation dataset. In addition, the expression of multiple immune cells was significantly elevated in PDs and IAs compared to controls, and both key crosstalk genes were significantly negatively associated with Macrophages M2. Finally, GATA2 was identified as a potential key transcription factor (TF), which regulates two key crosstalk genes and was upregulated in expression in the full dataset.

**Conclusion:** The present study identifies key crosstalk genes and TF in IAs and PD, providing new insights for further study of the co-pathogenesis of IAs and PD from an immune and inflammatory perspective.

## Introduction

Intracranial aneurysms (IAs) are a common neurosurgical condition and a major cause of nontraumatic subarachnoid hemorrhage^1^. The consequences of a ruptured intracranial aneurysm can be catastrophic. A study showed that the risk of death at 5, 10 and 15 years after subarachnoid hemorrhage was 12.9%, 23.6% and 35.4%, respectively^2^. Therefore, research into the pathogenesis of intracranial aneurysms, especially the prevention of unruptured intracranial aneurysms, is essential to avoid this type of hemorrhagic stroke.^3^.

Periodontitis (PD) is a chronic inflammation of the periodontal tissue caused mainly by local factors^4^, with an overall prevalence of 45%-50%.^5^. More and more evidence suggests that an association between IAs and PD. First, IAs share a high degree of risk factors with PD, including smoking and obesity.^6^. Second, investigations have shown that worsening periodontal disease parameters are significantly associated with the development of IAs and an increased risk of eventual aSAH^7^. The current view is that IAs are the end result of high-flow blood flow exerting high shear forces and flow-driven inflammatory cell-mediated remodeling of the intracranial arterial wall^8^. Up to 50% of IAs have DNA of oral pathogens in the aneurysmal wall^9^, such as Porphyromonasgingivalis. Porphyromonasgingivalis can remain viable in human macrophages and dendritic cells and may propagate to the aneurysmal wall of IAs through macrophage infiltration, which is strongly associated with inflammatory remodeling of IAs^10^. In addition, exposure to periodontal pathogens and immune response secondary to dysfunction may cause an increased risk of IAs formation and rupture in patients with periodontal disease^11^. These findings suggest a strong association between IAs and PD, but the specific molecular mechanisms and pathological interactions are not fully understood.

In this study, we tested and validated publicly available datasets from the GEO database. First, we identified differentially expressed genes (DEGs) of IAs and PD and obtained overlapping DEGs using the test datasets GSE54083 [IAs] and GSE10334 [PD]. Subsequently, we performed functional enrichment analysis on the overlapping DEGs. Next, we imported the overlapping DEGs into the STRING database to construct the PPI network and identified the PPI key genes using the cytoHubba plugin. WGCNA analysis were performed on the two test datasets separately and screened the gene modules most associated with IAs and PD disease traits. Crossover genes between PPI key genes and key modules were defined as key crosstalk genes. These key crosstalk genes were validated in the independent external datasets GSE75436 and GSE16134. In addition, we assessed the immune cell landscape of PDs and IAs by cibersort algorithm and further analyzed the correlation of key crosstalk genes with immune cells. Finally, this study also explored transcription factors (TFs) regulating key crosstalk genes, and finally obtained TFs commonly upregulated in IAs and PDs, which were further validated in the full dataset. Thus, our study may provide new clues to the co-pathogenesis of IAs and PD.

## Materials and Methods

### Data source

First we show the flow chart of this study (Figure 1). The expression data of IAs and PDs were obtained from the Gene Expression Omnibus data base (GEO) (https://www.ncbi.nlm.nih.gov/geo). The search strategy for this study included: (1) subject searches for “intracranial aneurysm” and “periodontitis”, respectively; (2) Study type option selection “Expression profiling by array”; (3) samples were obtained from Homo sapiens; (4) the dataset contained normal control group samples. The mRNA sequencing of the data set GSE54083 was based on GPL4133 Agilent-014850 Whole Human Genome Microarray 4×44K G4112F (Feature Number version). The mRNA sequencing of GSE10334 samples was based on GPL570 [HG-U133_Plus_2] Affymetrix Human Genome U133 Plus 2.0. The former includes 8 ruptured intracranial aneurysm samples, 5 unruptured aneurysm samples and 10 Superficial temporal artery samples, but the ruptured intracranial aneurysm samples will be excluded in this study. The latter contains 183 PD-affected gingival tissue samples and 64 unaffected gingival tissue samples.

**Fig. 1.**
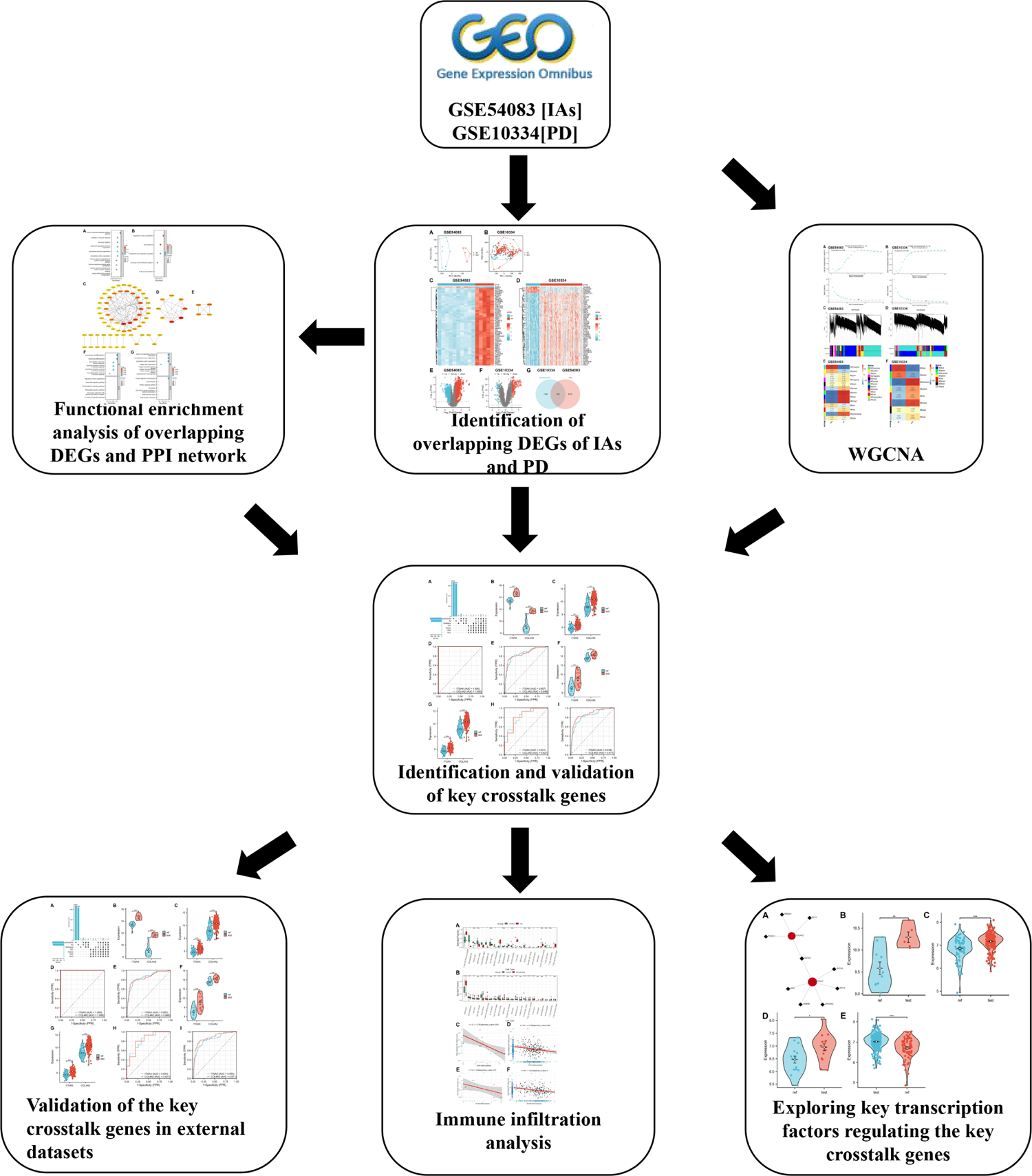
The flow diagram for the whole study

### Differentially expression analysis

Our preliminary analyses of the dataset were based on R software (version 4.2.1; https://cran.r-project.org). DEGs were identified by the “Limma” package^12^. The genes with |log2 fold change| > 0.5 and adjusted P-value < 0.05 were identified as DEGs. Genes that were both up-regulated or down-regulated in both sets of DEGs were defined as overlapping DEGs. Veen plots of overlapping DEGs were plotted with the help of the “ggvenn” package.

### Enrichment analysis of overlapping DEGs

We used the “clusterProfiler” package to perform enrichment analysis of overlapping DEGs, including Gene Ontology Biological Process (GO_BP) enrichment analysis^13^ We used the “clusterProfiler” package to perform enrichment analysis of overlapping DEGs, including GO_BP enrichment analysis.^14^ Terms with FDR < 0.05 were considered to be significantly enriched.

### Protein-protein interaction (PPI) network construction

We use the STRING database V11.5 (http://string-db.org) to build a PPI network with overlapping DEGs and set the “minimum required interaction score” parameter to 0.4 to hide the unconnected nodes. We obtained the interaction score of the overlapping DEGs, which was then imported the Cytoscape software V3.9.1^15^ for visualization. The MCODE plug in was used to filter clusters with high connectivity, thus dividing the PPI network into multiple clusters with default parameters. Finally, the genes in the clusters were analyzed for functional enrichment.

### Weighted gene co-expression network analysis

The top 5000 genes with the highest absolute median difference in expression in the test data set were screened for WGCNA analysis using the “WGCNA” package.^16^First, a soft threshold was obtained using the pickSoftThreshold function. Then, a weighted adjacency matrix was constructed. Finally, the correlation between each module and the disease was calculated. The module with the highest correlation with the grouped traits was defined as the key module. The genes within the key module were correlated with the grouped traits.

### Identification and validation of key crosstalk genes

First, we use the CytoHubba plugin of Cytoscape software to filter the hub genes in the PPI network^17^. The genes that were ranked in the top 20 using different algorithms (MCC, MNC, Degree and EPC) were identified as PPI key genes. Next, the PPI key genes were intersected with the key module genes of WGCNA, and these intersected genes were defined as key crosstalk genes. Subsequently, we compared the mRNA expression levels of key crosstalk genes between the case and control groups using the Mann-Whitney U test, and P < 0.05 was considered statistically significant, and visualized by “ggplot2”. Finally, we tested the diagnostic efficacy of key crosstalk genes in the test dataset using the receiver operating characteristic curves (ROCs) of the “pROC” package.

### Validation of key crosstalk genes in an independent external dataset

We validated the mRNA expression levels of key crosstalk genes in independent external datasets GSE75436 and GSE16134 samples. GSE75436 included 15 normal superficial temporal artery samples, 15 intracranial aneurysm samples, and mRNA sequencing of the samples was based on GPL570 [HG-U133_Plus_2] Affymetrix Human Genome U133 Plus 2.0 Array. GSE16134 contains 241 PD-affected gingival tissue samples and 69 unaffected gingival tissue samples, mRNA sequencing of the samples is based on GPL570 [HG-U133_Plus_2] Affymetrix Human Genome U133 Plus 2.0 Array.

### Immuno-infiltration analysis

First, immune cell expression levels in the test dataset were analyzed in the case and control groups using the “cibersort” algorithm^18^. We then calculated the correlation between the expression of key crosstalk genes and immune cell expression, P < 0.05 was considered statistically significant and was visualized by “ggplot2”.

### Identification of transcription factors (TFs) of key crosstalk genes

TFs of key crosstalk genes were predicted by NetworkAnalyst 3.0 (https://www.networkanalyst.ca)^19^. Subsequently, we compared the average expression levels of TFs in the test set and validation set samples using the Mann-Whitney U test. Finally, TFs that were typically upregulated in the case set were identified as potential key TFs in IAs and PDs.

## Results

### Identify overlapping DEGs of IAs and PDs

The “PCA” function was used to downscale the high-latitude data, and significant sample differences were found between the test and control groups in the two datasets (Figure 2A-D). In the dataset GSE54083, the “Limma” R package identified 4333 DEGs, of which 2364 were up-regulated and 1969 were down-regulated (Figure 2E). In the dataset GSE10334, the “Limma” R package identified 1353 DEGs, of which 813 were up-regulated and 540 were down-regulated (Figure 2F). Subsequently, we identified 258 intersecting DEGs (Figure 2G). Finally, the above genes were examined for trend checked and 127 overlapping DEGs were obtained (S1).

**Fig. 2.**
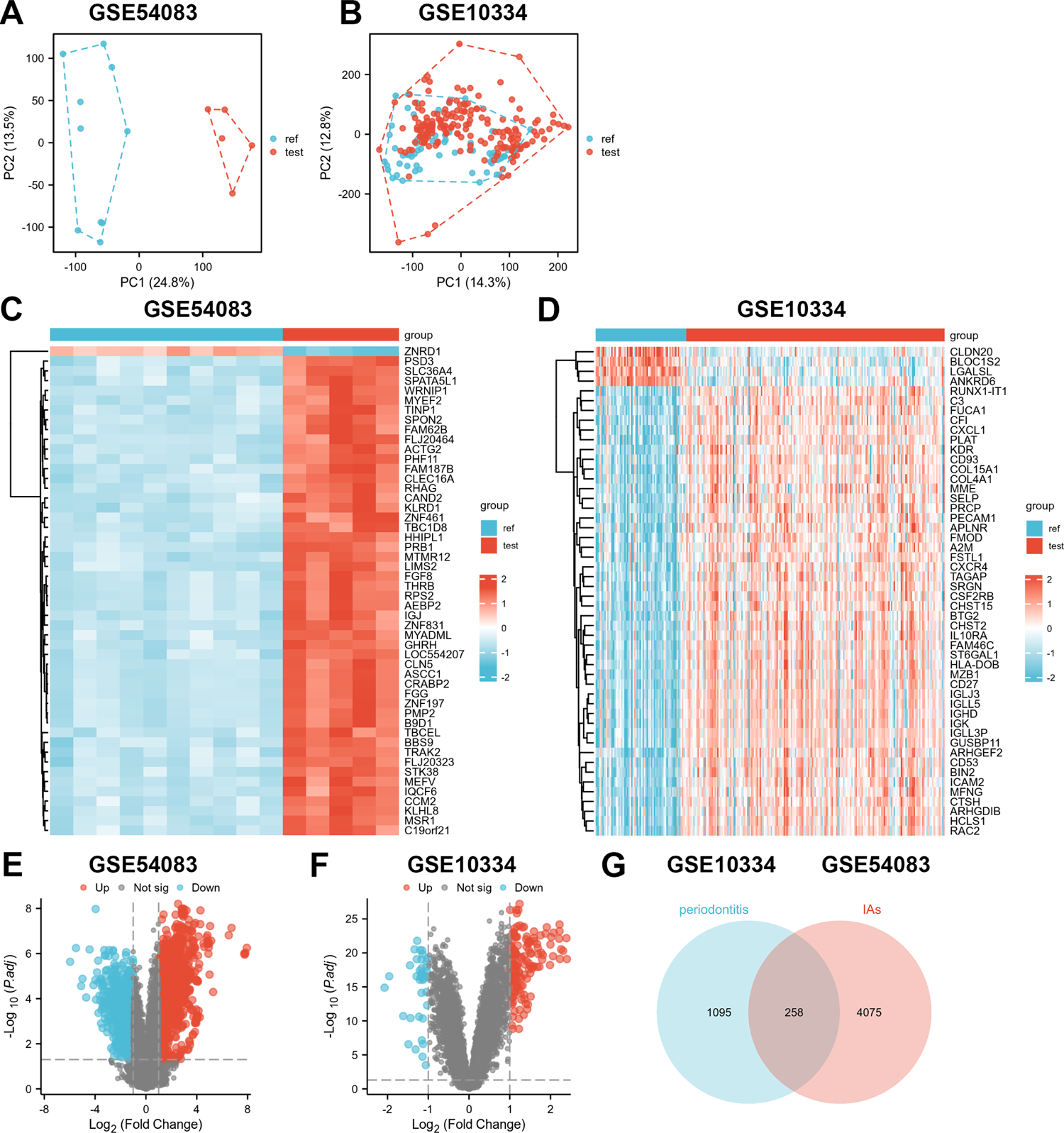
DEGs of test datasets GSE54083 and GSE10334.A, C, and E are PCA plots, heat maps, and volcano plots of GSE54083 DEG analysis, respectively. B, D, and F are PCA plots, heat maps, and volcano plots of GSE10334 DEG analysis, respectively. G. DEGs of GSE54083 and GSE10334 are crossed to obtain 258 intersecting DEGs.

### Functional enrichment analysis of overlapping DEGs

The GO_BP analysis showed that the most significantly enriched terms were immune response-regulating signaling pathway, extracellular matrix organization, extracellular structure organization, external encapsulating structure organization, leukocyte migration, and activation of immune response (Figure 3A). kEGG analysis showed that overlapping DEGs may be associated with Regulation of actin cytoskeleton, Axon guidance, Complement and coagulation cascades, and Leukocyte transendothelial migration (Figure 3B). Thus, overlapping DEG functions are clearly associated with immune and inflammatory processes.

**Fig. 3.**
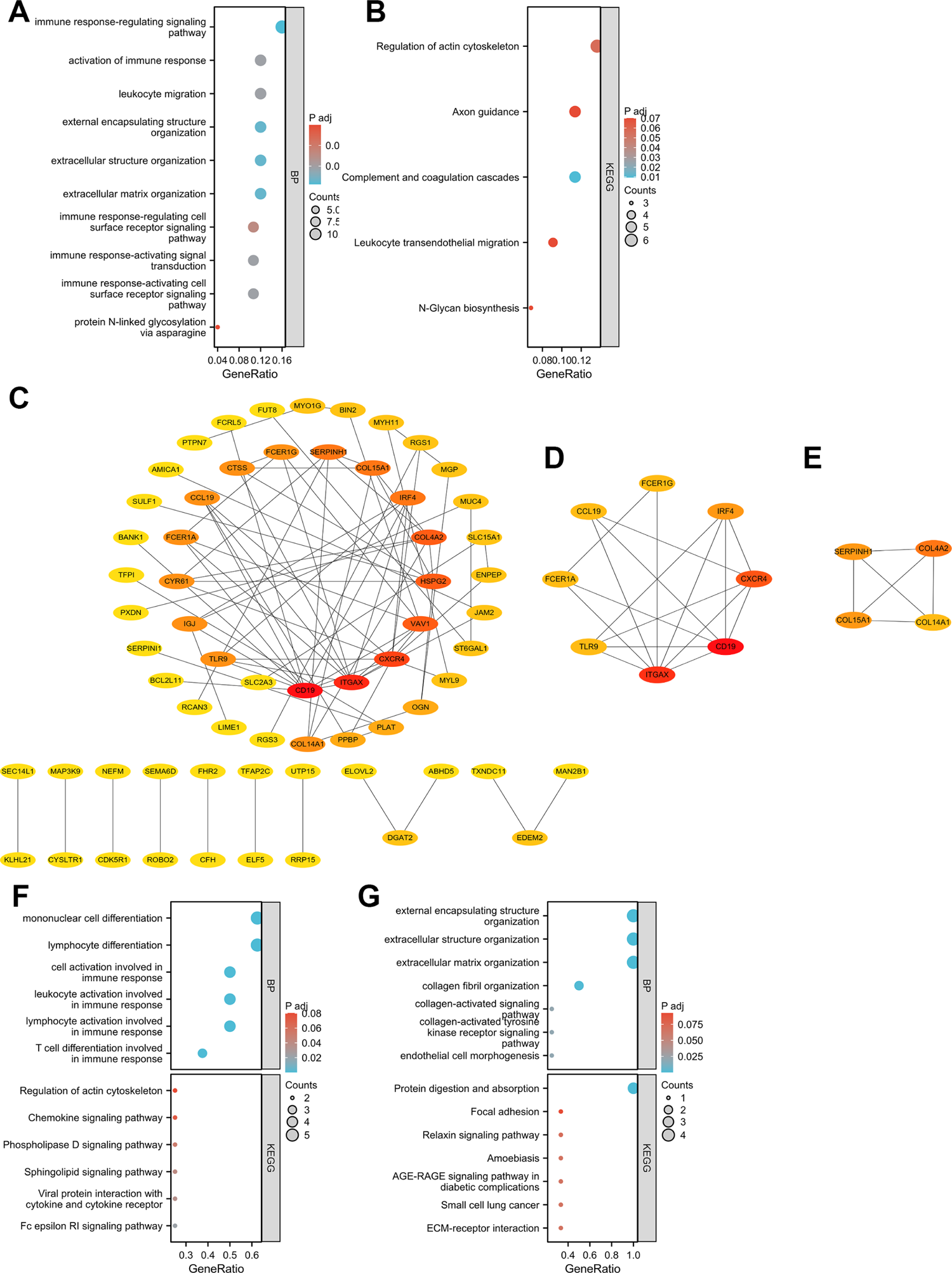
Functional enrichment analysis and PPI network construction for overlapping DEGs of the test dataset. A. GP-BP enrichment analysis of overlapping DEGs. B. KEGG enrichment analysis of overlapping DEGs. C. PPI network constructed for overlapping DEGs. D-E. PPI network obtained by the Mcode algorithm for cluster 1 and cluster 2. F-G. GP-BP enrichment analysis and KEGG enrichment analysis of genes contained in cluster 1 and cluster 2. enrichment analysis and KEGG enrichment analysis.

### Building PPI networks with overlapping DEGs

Overlapping DEGs were entered into the STRING database and a PPI network with 81 nodes and 123 edges (PPI enrichment p-value < 4.33e-09) was constructed at a medium confidence level (0.4). The PPI network was visualized using Cytoscape software (Figure 3C). Two clusters with high connectivity were identified by the Mcode plug-in (Figure 3D, E). Cluster 1 contains 8 nodes and 17 edges; Cluster 2 contains 4 nodes and 6 edges. Subsequently, we performed GP-BP enrichment analysis and KEGG enrichment analysis on the genes contained in Cluster 1 and Cluster 2 (Figure 3F, G). The functional enrichment of GO-BP for cluster 1 intronic genes mainly included lymphocyte differentiation, mononuclear cell differentiation, lymphocyte activation involved in immune response, leukocyte activation involved in immune response and T cell differentiation involved in immune response, etc. KEGG enrichment analysis for cluster 1 intronic gene mainly includes Fc epsilon RI signaling pathway, Viral protein interaction with cytokine and cytokine receptor and Sphingolipid signaling pathway, etc. The functional enrichment of GO-BP for cluster 2 intronic genes mainly included extracellular matrix organization, extracellular structure organization, external encapsulating structure organization, collagen fibril organization and collagen-activated signaling pathwa, etc. KEGG enrichment analysis for cluster 2 intronic genemainly included ECM-receptor interaction, AGE-RAGE signaling pathway in diabetic complications, Amoebiasis, Relaxin signaling pathway and Focal adhesion, etc.

### Weighted gene co-expression network construction and key module screening

The top 5000 genes with the highest absolute median difference in expression in the test dataset were screened for WGCNA analysis. β= 16 and β= 18 (scale-free R2 = 0.85) were chosen as soft thresholds for IAs and PDs, respectively, to ensure scale-free networks (Figure 4A and B). 13 modules were identified in GSE54083 and 8 modules in GSE10334 by WGCNA analysis (Figure 4C and D). Finally, a heat map of module-trait relationships was generated based on Pearson correlation coefficients. The results showed that the green module was the most correlated with IAs (0.854, p = 5.2E-05) and contained 661 genes; the turquoise module was the most correlated with PD (0.783, p = 5E-11) and included 705 genes (Figure 4E and F).

**Fig. 4.**
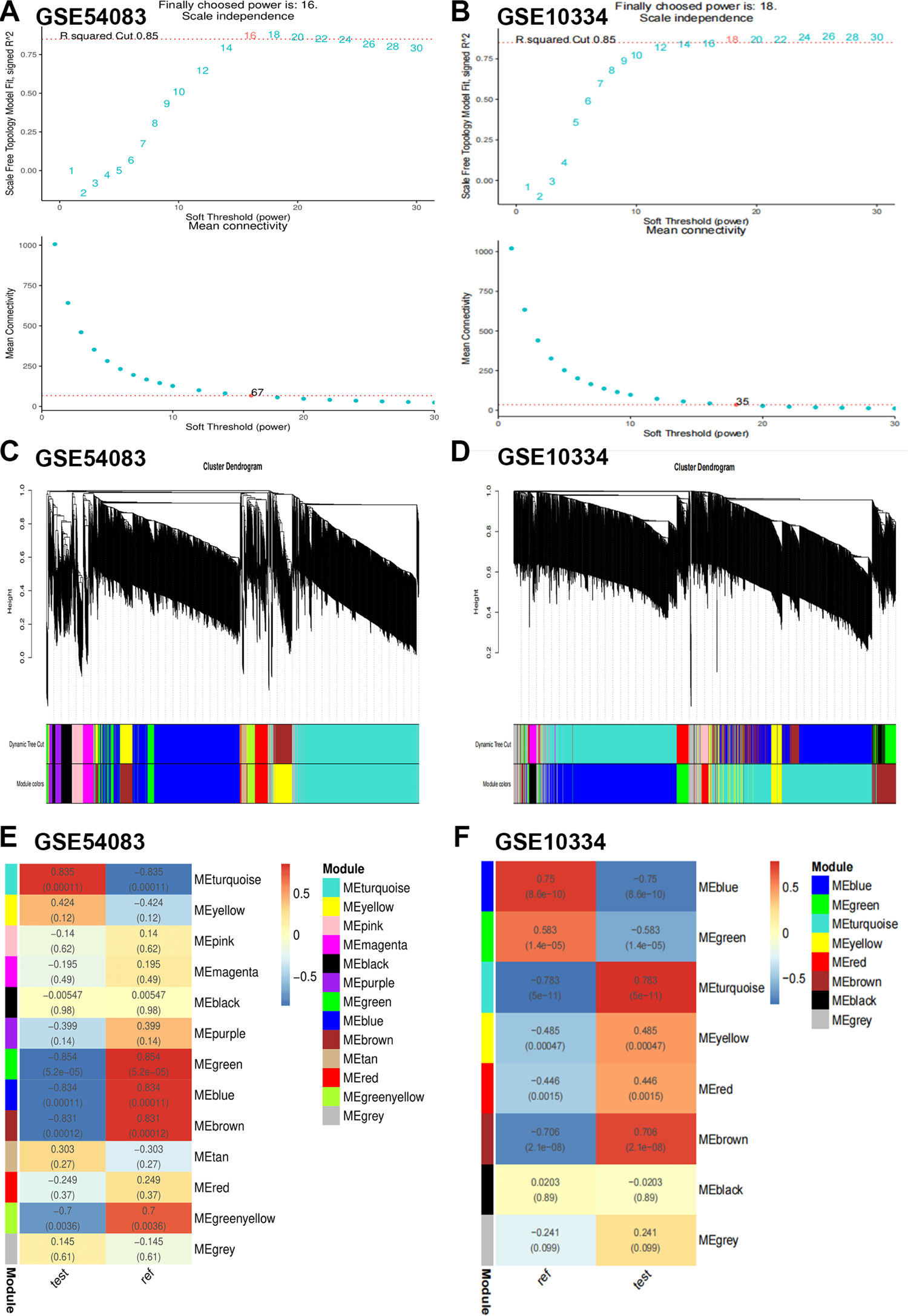
Weighted gene co-expression network analysis (WGCNA) of the test dataset. A-B. Soft threshold selection for GSE54083 and GSE10334. C-D. Clustering dendrogram of the top 5000 genes with the highest absolute median difference in expression between GSE54083 and GSE10334 based on differential measures. E-F. Module-trait relationship between GSE54083 and GSE10334 in relation to the modules and traits. Different colors represent different modules and contain the corresponding correlations and p-values.

### Identification and validation of key crosstalk genes

We used the CytoHubba plugin to sort the top 20 genes under four algorithms (Degree, EPC, MCC and MNC). The Upset plot showed that ITGAX and COL4A2 were present in both the top 20 genes of the four algorithms and in the two key modules (Figure 5A). Therefore, these two genes were identified as key crosstalk genes. In addition, the ITGAX and COL4A2 mRNA expression levels were significantly higher in the case group than in the control group in GSE54083 and GSE10334 (Figure 5B, C). In addition, ITGAX and COL4A2 had good diagnostic ability for IAs and PD according to ROC curves (Figure 5D, E).

**Fig. 5.**
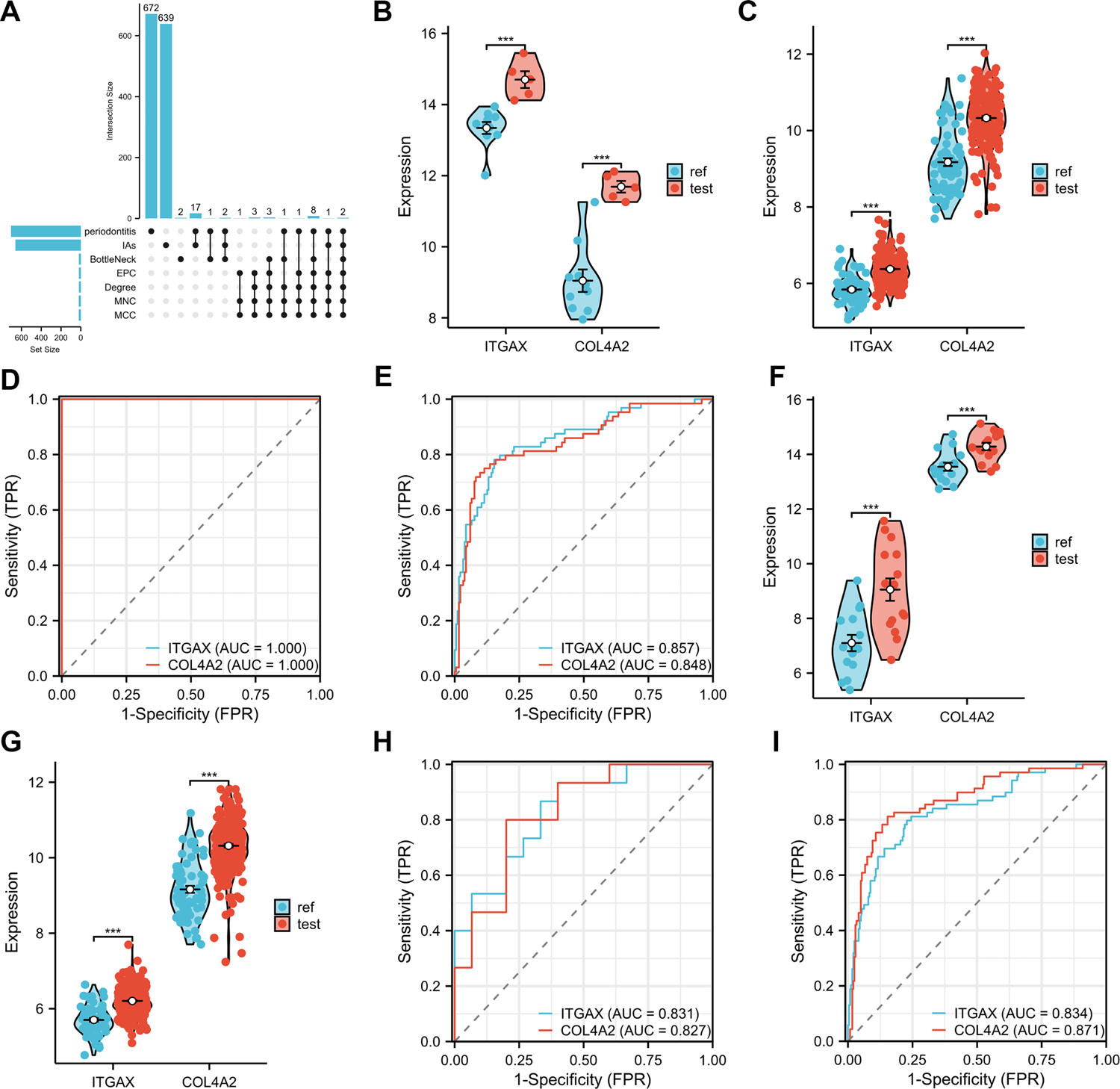
Identification and validation of key crosstalk genes. A. ITGAX and COL4A2 were both present in the four algorithms and two WGCNA key modules. B-C. ITGAX and COL4A2 expression levels were higher in the case group than in the control group in GSE54083 and GSE10334. D-E. ITGAX and COL4A2 had good diagnostic capabilities in GSE54083 and GSE10334. F-G. The expression levels of ITGAX and COL4A2 were higher in the GSE75436 and GSE16134 case groups than in the control group. H-I. ITGAX and COL4A2 had good diagnostic capabilities in GSE75436 and GSE16134. (***, p < 0.0001).

### Validation of key crosstalk genes in an independent external dataset

To increase the confidence level, we validated the expression of key crosstalk genes in two additional independent external datasets (GSE75436 and GSE16134). Consistent with the results of the test set, ITGAX and COL4A2 mRNA expression levels were elevated in the case set (Figure 5F, G), while showing good diagnostic efficiency (Figure 5H, I).

### Immuno-infiltration analysis

By performing enrichment analysis of overlapping DEGs, we found that immune and inflammatory processes are involved in crosstalk between IAs and PD. Therefore, we used the cibersort algorithm to analyze the proportion of immune cells in the case group versus control samples in both test datasets. We found that the immune landscape of IAs and PD was significantly altered in the case group (Figure 6A and B). Furthermore, correlation analysis showed that ITGAX and COL4A2 expression levels were significantly negatively correlated with Macrophages M2 (Figure 6C-F).

**Fig. 6.**
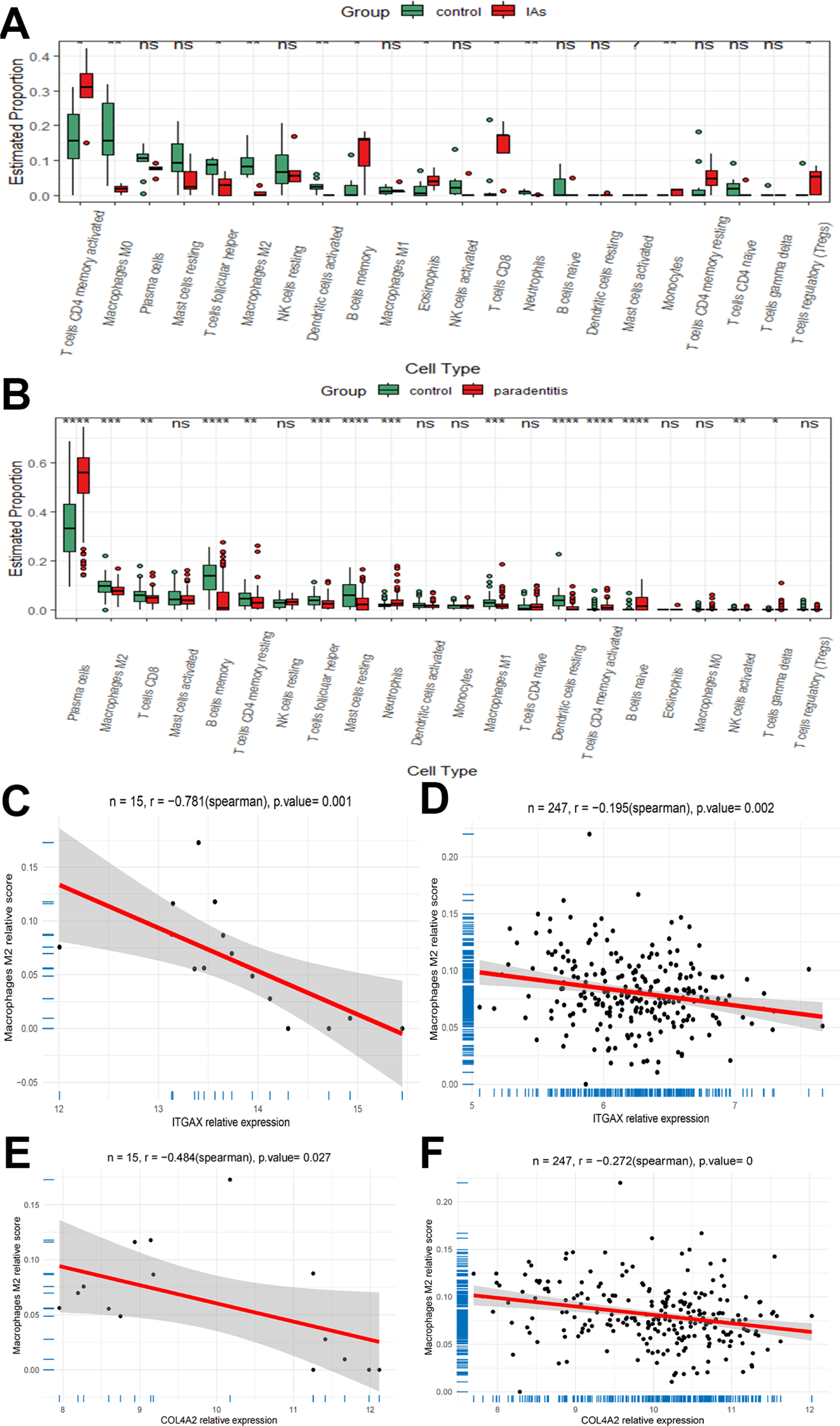
Immune infiltration analysis of the test dataset. A-B. The immune landscape was significantly altered in the GSE54083 and GSE10334 case groups. C-D. ITGAX and COL4A2 expression levels in GSE54083 were significantly negatively correlated with Macrophages M2. D-F. ITGAX and COL4A2 expression levels in GSE10334 were significantly negatively correlated with Macrophages M2.

### Exploring key transcription factors (TFs) that regulate key crosstalk genes

Further, we explored potential TFs that may regulate ITGAX and COL4A2 genes using NetworkAnalyst 3.0 and compared the expression levels of case and control groups in all datasets by Mann-Whitney U test. We found that GATA2 interacted with both ITGAX and COL4A2 (Figure 7A), and the expression level of GATA2 was significantly elevated in all case groups (Figure 7B). Thus, GATA2 may be a potential key TF regulating two key crosstalk genes in the pathological process of IAs and PD.

**Fig. 7.**
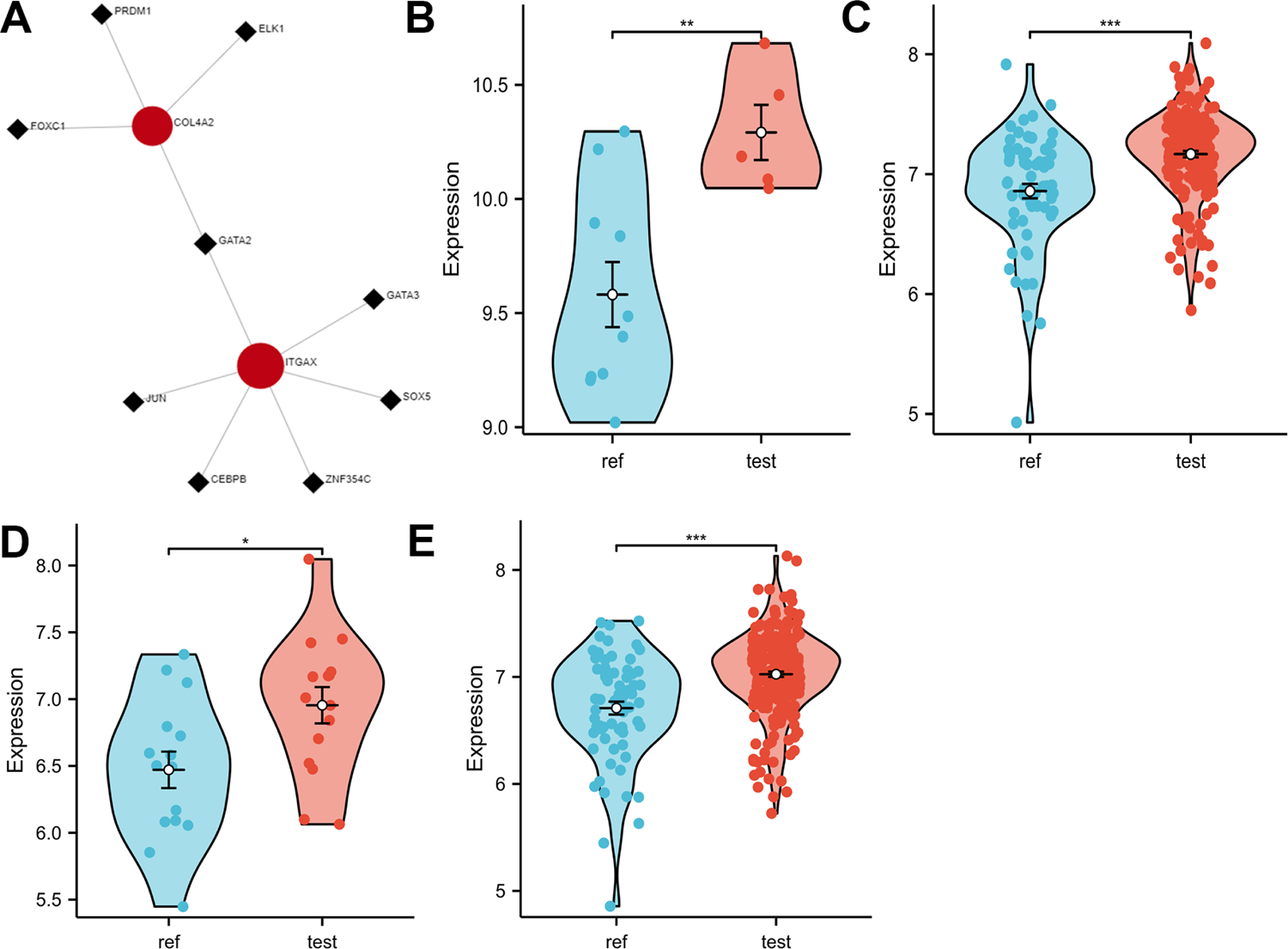
GATA2 was identified as a potential key TF shared in IAs and PD. A. NetworkAnalyst 3.0 suggests that GATA2 regulates both ITGAX and COL4A2. B-C. GATA2 was significantly elevated in the test dataset (GSE54083 and GSE10334) case set. D-E. GATA2 was significantly elevated in the external validation dataset (GSE75436 and GSE16134) case group (*, P < 0.01; **, P < 0.001; ***, P < 0.0001).

## Discussion

Current studies suggest that periodontitis is not only associated with abdominal aortic aneurysm^20^, coronary heart disease^21^ and atherosclerosis^22^ cardiovascular disease risk, but also with the occurrence and rupture of intracranial aneurysms.^11, 23^ However, the specific mechanisms of the effect of periodontitis on intracranial aneurysms have not been well studied. In the present study, we focused on exploring the key crosstalk genes (ITGAX and COL4A2) shared by intracranial aneurysms and periodontitis as well as the transcription factor (GATA2) regulating key crosstalk genes by bioinformatics approach, which was fully validated in an external independent dataset. To our knowledge, this is the first study to report the above findings.

Through a comprehensive analysis of the dataset, ITGAX and COL4A2 were identified as key crosstalk genes in the comorbidity of IAs and PD. ITGAX, also known as CD11c, complement receptor 4 (CR4), is responsible for encoding integrin αX chain proteins, a marker shared by macrophages and dendritic cells.^24^ ITGAX can promotes monocyte adhesion and chemotaxis, regulates immune response, and plays an important role in the pathogenesis of infection and atherosclerosis.^25^. Macrophages and the inflammatory signals they mediate play an important role in the pathogenesis of IAs^26, 27^. Meanwhile, macrophage levels were significantly higher in periodontitis samples than in healthy samples^28^, they are central players in the destruction and repair phases of periodontal disease.^29^ COL4A2, which encodes the α2 chain of type IV collagen, is a major structural component of basement membranes (BMs) and plays a fundamental and crucial role in vessel wall integrity; Abnormal COL4A2 expression can lead to a broad phenotypic spectrum involving the nervous system, kidneys, and other organs, but the main site of vascular damage is the brain^30, 31^. Compared to familial intracranial aneurysms, abnormal COL4A2 expression in sporadic aneurysms may be associated with aneurysm development ^32, 33^. Periodontitis is characterized by irreversible and progressive degeneration of periodontal tissue. Periodontal ligament stem cells (PDLSC) were involve in periodontal tissue regeneration, and COL4A2 in the tissue-specific extracellular matrix plays an important role in this process^34^. Therefore, two upregulated key crosstalk genes may be involved in IAs and PD comorbidity mechanisms.

The role of immune infiltration in IAs is of increasing interest. Crosstalk gene enrichment analysis suggests that immune and inflammatory processes are involved in IAs and PD comorbidity mechanisms. Immune cell infiltration analysis revealed a significantly different immune landscape in IAs and PD disease groups compared to controls, and two key crosstalk genes expression was significantly negatively correlated with Macrophages M2. Previous studies have shown that macrophage-mediated cellular and molecular inflammation is a key factor in aneurysm formation and rupture^35^. The inflammatory process upregulated by M1 macrophages underlies the development of intracranial aneurysms, and a balanced shift to M2 prevents aneurysm formation^36^. Similarly, M1 polarization is a major feature of macrophages in periodontitis and M2 differentiation is significantly inhibited^37^, which may contribute to the development and progression of periodontal tissue destruction in periodontitis^29^. These findings are consistent with the results of the present study. Therefore, these two key crosstalk genes may promote IAs and PD comorbidity through immunity and inflammation.

To investigate the candidate regulatory mechanisms of key crosstalk genes (ITGAX and COL4A2) shared by intracranial aneurysms and periodontitis, we constructed a target gene-TF network. Our study showed that the potential regulatory network of ITGAX and COL4A2 consists of 9 TFs, in which GATA2 interacts closely with both of these genes. Recent studies have shown that GATA2 may play an important role in regulating the immune response in the mechanism of IAs formation^38^. Meanwhile, phagocytic GATA2 overexpression drives atherosclerosis formation.^39, 40^. These findings are consistent with the results of the present study. Currently, no studies have focused on the potential role of GATA2 in the pathogenesis of PD. According to the available data, GATA2 is a zinc-finger transcription factor that is mainly expressed in the hematopoietic system.GATA2 regulates various biological processes to directly or directly influence the progression of atherosclerosis, including aortic neovascularization, hematopoiesis, adipogenesis, and inflammation^41, 42^. For example, it improves monocyte adhesion and promotes leakage through the arterial wall by upregulating VCAM-1^42^.

This study identifies key crosstalk genes and TFs in IAs and PD from an immune and inflammatory perspective, thus providing new insights into the mechanisms of comorbidity in IAs and PD. It is important to note that the conclusion needs more clinical validation in the future. In addition, the specific functions of the crosstalk genes remain to be validated in vivo and in vitro.

## Conclusion

In summary, we identified ITGAX and COL4A2 as key crosstalk genes in IAs and PD by multiple bioinformatics analysis methods, and they may be involved in crosstalk between IAs and PD through immune pathways. In addition, GATA2 was identified as a potential key TF in IAs and PD. To our knowledge, this is the first study to report the above findings. The present study provides new insights into the co-pathogenesis of IAs and PD, and the current results require further validation.

## Conflict of Interest

The authors declare that the research was conducted in the absence of any commercial or financial relationships that could be construed as a potential conflict of interest.

## Authors’ contributions

Jianhuang Huang analyzed the data and wrote this manuscript. Yao Chen, Yuanbao Kang and Jianhuang Song assisted in analyzing the data and revising the manuscript. Zhengjian Yao critically read and edited the manuscript.

## Data Availability

The data used to support the findings of this study are available from the corresponding author upon request

https://www.jianguoyun.com/p/DZACmZYQhKCkCRj_gYkFIAA

## Acknowledgements

Not applicable.

## Abbreviations

IAs: Intracranial Aneurysm

PD: Periodontitis

GEO: Gene Expression Omnibus

DEG: Differentially expressed gene

PPI: Protein-protein Interaction

WGCNA: Weighted Gene Co-expression Network Analysis

TF: Transcription factor.

## Availability of data and materials

The datasets analyzed in this study are available for download in the Gene Expression Omnibus (GEO) database,https://www.ncbi.nlm.nih.gov/geo

## Notes

### Competing Interest Statement

The authors have declared no competing interest.

### Funding Statement

none

### Author Declarations

Affiliated Hospital of Putian University

## References

1. Alshekhlee A, Mehta S, Edgell RC, Vora N, Feen E, Mohammadi A, et al. Hospital mortality and complications of electively clipped or coiled unruptured intracranial aneurysm. Stroke. 2010;41:1471–1476

2. Sharma M, Brown B, Madhugiri V, Cuellar-Saenz H, Sonig A, Ambekar S, et al. Unruptured intracranial aneurysms: Comparison of perioperative complications, discharge disposition, outcome, and effect of calcification, between clipping and coiling: A single institution experience. Neurol India. 2013;61:270–276

3. van Gijn J, Kerr RS, Rinkel GJ. Subarachnoid haemorrhage. Lancet. 2007;369:306–318

4. Papapanou PN, Sanz M, Buduneli N, Dietrich T, Feres M, Fine DH, et al. Periodontitis: Consensus report of workgroup 2 of the 2017 world workshop on the classification of periodontal and peri-implant diseases and conditions. J Periodontol. 2018;89 Suppl 1:S173–s182

5. Kassebaum NJ, Bernabé E, Dahiya M, Bhandari B, Murray CJ, Marcenes W. Global burden of severe periodontitis in 1990-2010: A systematic review and meta-regression. J Dent Res. 2014;93:1045–1053

6. Hallikainen J, Keränen S, Savolainen J, Närhi M, Suominen AL, Ylöstalo P, et al. Role of oral pathogens in the pathogenesis of intracranial aneurysm: Review of existing evidence and potential mechanisms. Neurosurg Rev. 2021;44:239–247

7. Pyysalo MJ, Pyysalo LM, Pessi T, Karhunen PJ, Öhman JE. The connection between ruptured cerebral aneurysms and odontogenic bacteria. J Neurol Neurosurg Psychiatry. 2013;84:1214–1218

8. Leite FRM, Nascimento GG, Scheutz F, López R. Effect of smoking on periodontitis: A systematic review and meta-regression. Am J Prev Med. 2018;54:831–841

9. Beukers NG, van der Heijden GJ, van Wijk AJ, Loos BG. Periodontitis is an independent risk indicator for atherosclerotic cardiovascular diseases among 60174 participants in a large dental school in the netherlands. J Epidemiol Community Health. 2017;71:37–42

10. Hu S, Yu N, Li Y, Hao Z, Liu Z, Li MH. A meta-analysis of risk factors for the formation of de novo intracranial aneurysms. Neurosurgery. 2019;85:454–465

11. Liu K, Sun J, Shao L, He H, Liu Q, Li Y, et al. Correlation of periodontal diseases with intracranial aneurysm formation: Novel predictive indicators. Chin Neurosurg J. 2021;7:31

12. Ritchie ME, Phipson B, Wu D, Hu Y, Law CW, Shi W, et al. Limma powers differential expression analyses for rna-sequencing and microarray studies. Nucleic Acids Res. 2015;43:e47

13. Ashburner M, Ball CA, Blake JA, Botstein D, Butler H, Cherry JM, et al. Gene ontology: Tool for the unification of biology. The gene ontology consortium. Nat Genet. 2000;25:25–29

14. Kanehisa M, Goto S. Kegg: Kyoto encyclopedia of genes and genomes. Nucleic Acids Res. 2000;28:27–30

15. Shannon P, Markiel A, Ozier O, Baliga NS, Wang JT, Ramage D, et al. Cytoscape: A software environment for integrated models of biomolecular interaction networks. Genome Res. 2003;13:2498–2504

16. Langfelder P, Horvath S. Wgcna: An r package for weighted correlation network analysis. BMC Bioinformatics. 2008;9:559

17. Chin CH, Chen SH, Wu HH, Ho CW, Ko MT, Lin CY. Cytohubba: Identifying hub objects and sub-networks from complex interactome. BMC Syst Biol. 2014;8 Suppl 4:S11

18. Newman AM, Liu CL, Green MR, Gentles AJ, Feng W, Xu Y, et al. Robust enumeration of cell subsets from tissue expression profiles. Nat Methods. 2015;12:453–457

19. Zhou G, Soufan O, Ewald J, Hancock REW, Basu N, Xia J. Networkanalyst 3.0: A visual analytics platform for comprehensive gene expression profiling and meta-analysis. Nucleic Acids Res. 2019;47:W234–w241

20. Salhi L, Sakalihasan N, Okroglic AG, Labropoulos N, Seidel L, Albert A, et al. Further evidence on the relationship between abdominal aortic aneurysm and periodontitis: A cross-sectional study. J Periodontol. 2020;91:1453–1464

21. Nocini R, Favaloro EJ, Sanchis-Gomar F, Lippi G. Periodontitis, coronary heart disease and myocardial infarction: Treat one, benefit all. Blood Coagul Fibrinolysis. 2020;31:339–345

22. Kebschull M, Demmer RT, Papapanou PN. “Gum bug, leave my heart alone!”--epidemiologic and mechanistic evidence linking periodontal infections and atherosclerosis. J Dent Res. 2010;89:879–902

23. Hallikainen J, Lindgren A, Savolainen J, Selander T, Jula A, Närhi M, et al. Periodontitis and gingival bleeding associate with intracranial aneurysms and risk of aneurysmal subarachnoid hemorrhage. Neurosurg Rev. 2020;43:669–679

24. Liu P, Yu YR, Spencer JA, Johnson AE, Vallanat CT, Fong AM, et al. Cx3cr1 deficiency impairs dendritic cell accumulation in arterial intima and reduces atherosclerotic burden. Arterioscler Thromb Vasc Biol. 2008;28:243–250

25. Cole JE, Park I, Ahern DJ, Kassiteridi C, Danso Abeam D, Goddard ME, et al. Immune cell census in murine atherosclerosis: Cytometry by time of flight illuminates vascular myeloid cell diversity. Cardiovasc Res. 2018;114:1360–1371

26. Shimizu K, Kushamae M, Mizutani T, Aoki T. Intracranial aneurysm as a macrophage-mediated inflammatory disease. Neurol Med Chir (Tokyo). 2019;59:126–132

27. Stratilová MH, Koblížek M, Štekláčová A, Beneš V, Sameš M, Hejčl A, et al. Increased macrophage m2/m1 ratio is associated with intracranial aneurysm rupture. Acta Neurochir (Wien). 2023;165:177–186

28. Garaicoa-Pazmino C, Fretwurst T, Squarize CH, Berglundh T, Giannobile WV, Larsson L, et al. Characterization of macrophage polarization in periodontal disease. J Clin Periodontol. 2019;46:830–839

29. Zhou LN, Bi CS, Gao LN, An Y, Chen F, Chen FM. Macrophage polarization in human gingival tissue in response to periodontal disease. Oral Dis. 2019;25:265–273

30. Volonghi I, Pezzini A, Del Zotto E, Giossi A, Costa P, Ferrari D, et al. Role of col4a1 in basement-membrane integrity and cerebral small-vessel disease. The col4a1 stroke syndrome. Curr Med Chem. 2010;17:1317–1324

31. Meuwissen ME, Halley DJ, Smit LS, Lequin MH, Cobben JM, de Coo R, et al. The expanding phenotype of col4a1 and col4a2 mutations: Clinical data on 13 newly identified families and a review of the literature. Genet Med. 2015;17:843–853

32. Liu Y, Song Y, Liu P, Li S, Shi Y, Yu G, et al. Comparative bioinformatics analysis between proteomes of rabbit aneurysm model and human intracranial aneurysm with label-free quantitative proteomics. CNS Neurosci Ther. 2021;27:101–112

33. Mohan D, Munteanu V, Coman T, Ciurea AV. Genetic factors involves in intracranial aneurysms--actualities. J Med Life. 2015;8:336–341

34. Wen Y, Yang H, Wu J, Wang A, Chen X, Hu S, et al. Col4a2 in the tissue-specific extracellular matrix plays important role on osteogenic differentiation of periodontal ligament stem cells. Theranostics. 2019;9:4265–4286

35. Muhammad S, Chaudhry SR, Dobreva G, Lawton MT, Niemelä M, Hänggi D. Vascular macrophages as therapeutic targets to treat intracranial aneurysms. Front Immunol. 2021;12:630381

36. Nowicki KW, Hosaka K, Walch FJ, Scott EW, Hoh BL. M1 macrophages are required for murine cerebral aneurysm formation. J Neurointerv Surg. 2018;10:93–97

37. Almubarak A, Tanagala KKK, Papapanou PN, Lalla E, Momen-Heravi F. Disruption of monocyte and macrophage homeostasis in periodontitis. Front Immunol. 2020;11:330

38. Li S, Xiao J, Yu Z, Li J, Shang H, Zhang L. Integrated analysis of c3ar1 and cd163 associated with immune infiltration in intracranial aneurysms pathogenesis. Heliyon. 2023;9:e14470

39. Yin C, Vrieze AM, Rosoga M, Akingbasote J, Pawlak EN, Jacob RA, et al. Efferocytic defects in early atherosclerosis are driven by gata2 overexpression in macrophages. Front Immunol. 2020;11:594136

40. Luo S, Wang F, Chen S, Chen A, Wang Z, Gao X, et al. Nrp2 promotes atherosclerosis by upregulating parp1 expression and enhancing low shear stress-induced endothelial cell apoptosis. Faseb j. 2022;36:e22079

41. Izadpanah P, Khabbzi E, Erfanian S, Jafaripour S, Shojaie M. Case-control study on the association between the gata2 gene and premature myocardial infarction in the iranian population. Herz. 2021;46:71–75

42. Luo C, Tang B, Qin S, Yuan C, Du Y, Yang J. Gata2 regulates the cad susceptibility gene adtrp rs6903956 through preferential interaction with the g allele. Mol Genet Genomics. 2021;296:799–808

